# Low-Dose Naltrexone use for the management of post-acute sequelae of COVID-19

**DOI:** 10.1101/2023.06.08.23291102

**Authors:** Hector Bonilla, Lu Tian, Vincent C. Marconi, Robert Shafer, Grace A. McComsey, Mitchel Miglis, Philip Yang, Linda N. Geng

## Abstract

Post-Acute Sequelae of SARS-CoV-2 (PASC), also known as Long COVID, is globally estimated to have affected up to 40-50% of individuals who were infected with SARS-CoV-2. The causes of PASC are being investigated, and there are no established therapies. One of the leading hypotheses for the cause of PASC is the persistent activation of innate immune cells with increased systemic inflammation. Naltrexone is a medication with anti-inflammatory and immunomodulatory properties that has been used in other conditions that overlap with PASC. In this study we performed retrospective review of a clinical cohort of 59 patients at a single academic center who received low-dose naltrexone (LDN) off-label as a potential therapeutic intervention for PASC. The use of LDN was associated with improved clinical symptoms (fatigue, brain fog, post exertional malaise/PEM, unrefreshing sleep, sleep pattern, and headache), fewer number of symptoms, and better functional status. This observational finding warrants further testing in rigorous, randomized, placebo-controlled clinical trials.

**Highlights:**

- The off-label use of low-dose naltrexone is a promising drug intervention candidate for the management of post-COVID conditions.
- Low-dose naltrexone has anti-inflammatory and immunomodulatory properties which may benefit those with PASC where persistent inflammation is the causative pathway.
- This potential benefit of LDN warrants further testing in rigorous randomized, placebo-controlled clinical trials.

## Introduction

The NIH has provided guidelines on treating acute COVID-19 for hospitalized and non-hospitalized adults and children. However, recommendations are lacking for patients with Long COVID, also known as post-acute sequelae of SARS-CoV-2 (PASC) (1). In September 2021, the NIH launched the Researching COVID to Enhance Recovery (RECOVER) Initiative to understand the clinical manifestations and pathobiology of post-COVID conditions and test interventions for patients with PASC (2). One of the leading hypotheses of PASC pathophysiology is immune and inflammatory dysregulation and, therefore, the therapeutic use of immune modulators and anti-inflammatory medications deserves investigation (3, 4).

Naltrexone hydrochloride is an oral μ-opioid receptor antagonist that has immune-modulating properties. Its first-pass metabolism ranges from 5 to 40% and therapeutic activity is due to naltrexone and its 6-ß-naltrexol metabolite; both are renally excreted (53% to 79% of the dose). The half-life for naltrexone and 6-ß-naltrexol is 4 hours and 13 hours, respectively. Off-label low-dose naltrexone (LDN) has been used for fibromyalgia, Crohn’s disease, multiple sclerosis, complex regional pain syndrome, Hailey-Haile disease, cancer, myalgic encephalomyelitis/chronic fatigue syndrome (ME/CFS), and PASC (5-8). There is a significant overlap between ME/CFS and PASC symptoms (9). In a cohort of 140 patients with PASC, 46% also met the diagnostic criteria for ME/CFS (10). Here, we report patients’ experience at a single academic PASC clinic who received Low-dose Naltrexone (LDN) off-label as a potential therapeutic intervention for PASC.

## Material and Methods

This report employed a detailed retrospective review of Electronic Health Records (EHR) from 468 adult patients in the Stanford PASC clinic with a history of SARS-CoV-2 infection and symptoms attributable to PASC (9). We reviewed the charts for patients who received LDN off-label for the indication of PASC between May 18, 2021, to March 17, 2023. Low-dose naltrexone was typically prescribed as individualized dose-titration ranging from 0.5 mg daily to 6.0 mg daily. All patients had a documented previous positive test for SARS-CoV-2 (PCR, antigen detection, or a positive serology before SARS-CoV-2 vaccination), and persistent symptoms for ≥28 days following infection. In addition, symptoms were ascertained using a detailed questionnaire that was completed by patients within seven days before each clinic visit. The questionnaire included (i) the 29 symptoms commonly reported to occur in patients with acute COVID-19; (ii) the severity of each symptom based on the Likert scale (mild=1, moderate=2, severe=3 very severe=4 and incapacitating=5) (iii) the Post COVID-19 Functional Status Scale (FSS) modified from Klok et al. (11), which classifies patients as either asymptomatic (level I), symptomatic without limitations (level II), symptomatic with reduced daily activity (level III), symptomatic with a struggle to perform daily activities (level IV), or incapacitated and bedridden (level V). Out of the 29 symptoms captured, the seven most frequent symptoms identified from our PASC cohort were selected for our primary comparative pre- and post-treatment analyses: headache, fatigue, brain fog, unrefreshing sleep, sleep difficulties, post-exertional malaise, and lightheadedness. We used Research Electronic Data Capture (REDCap) and Excel platforms for data collection. The current study is part of IRB Stanford Post-Acute COVID-19 Syndromes: Patient Database, which received approval from the Stanford Institutional Review Board; protocol #62996. Any SARS-CoV-2 infection before December 1, 2021, was considered an infection caused by a non-Omicron variant.

Out of 468 patients seen in the clinic during the pre-specified time points, LDN was prescribed for 222 patients; among these, 59 patients who had completed the questionaries at every clinic visit were included in the analysis (supplementary data whole data).

### Statistical Analysis

Patient characteristics were compared between clinical symptoms using the Mann-Whiney rank sum test and summarized using median and interquartile by response status. Similarly, we compared the patient characteristics and responses by the time since initial COVID-19 infections (<365 days vs ≥ 365 days). The number of symptoms before and after LDN was summarized and compared using the Wilcoxon rank sum test. Statistically significant improvement was defined by (P value<0.05). The correlation coefficient was calculated between the LDN dose and clinical symptom scores.

## Results

Among the 59 PASC patients from our clinic who received LDN and completed questionnaires, the median age was 45 years (IQR 34-59), and the majority were women (n=40; 67.8%). The median duration of symptoms from initial COVID-19 infection to LDN use was 361 days (range 61-708 days), the median baseline number of symptoms was 14 (IQR 9-16), and the functional status scale was Level II in 1, level III in 23 patients, Level IV in 29, and level V in 6 patients. The majority of the SARS-CoV-2 infections (83.3%) occurred in the pre-Omicron era, and 83.3% of patients had at least one dose of the COVID vaccine before infection.

At the last office visit, the LDN dose ranged from 0.5 to 6 mg; the median dose was 2.0 mg [IQR 1-5]. 62.7% (n=37) patients reported a subjective improvement in at least 1 of their symptoms after a minimum of 31 days of treatment (median 143 days). The patients reported subjective improvement in fatigue, brain fog, sleep, pain, and others (headache, anxiety, breathing, stuttering, and voice strength). Five patients reported severe side effects including: “nightmares,” “feeling poisoned,” and “worsening symptoms” that led to discontinuation of the drug. The median days on LDN was 143 days [IQR 77-255].

Although some people with PASC symptoms can experience symptom resolution within a few weeks to months after infection, the median duration of illness in our study cohort was 358 days from the initial infection. Symptom responses were stratified by the duration by comparing patients with symptoms ≥ 365 days (one year) against patients with symptoms <365 days. Twenty-eight patients had symptoms for <365 days (47.5%), and 31 had symptoms for ≥365 days (52.5%). There was no statistical difference between these two groups regarding age, days on LDN, number of symptoms, changes in the number of symptoms, or improvement in the selected symptoms. (Table 2.)

**Table 1.** Characteristics and symptom changes in patients with PASC receiving Low Dose Naltrexone (0.5-6 mg) by the duration of infection.

|  | **>=365 days** | **<365 days** | **P-value** |
| --- | --- | --- | --- |
| Cases | 28 (47.5%) | 31 (52.5%) |  |
| Number of symptoms at baseline | 14 [9, 16]* | 14 [8.5, 17] | 0.749 |
| Days since infection | 496 [452, 593] | 268 [131, 326] |  |
| Age | 45 [36, 55] | 48 [34, 60] | 0.885 |
| Female | 21(75%) | 19(61.3%) | 0.282 |
| Days on LDN** | 133 [75, 179] | 143 [86, 319] | 0.179 |
| Change in number of symptoms | -1.5 [-3, 0] | -2 [-5.5, 0.5] | 0.393 |
| Change in Fatigue | -1 [-1, 0] | 0 [-2, 0.5] | 0.608 |
| Change in Brain fog | 0 [-1,0.25] | 0 [-1, 0] | 0.788 |
| Change in Post exertional malaise *** | -0.5 [-1, 0] | 0 [-1.5, 0] | 1.000 |
| Change in Unrefreshing sleep | 0 [-2, 0] | 0 [-1.5, 0.5] | 0.297 |
| Change Abnormal sleep pattern | -1 [-2.25, 0] | 0 [-1.5, 0] | 0.655 |
| Change in Headache | -0.5 [-1.25, 0] | 0 [-1, 1] | 0.462 |
| Change in Lightheadedness | 0 [0, 0.25] | 0 [-1, 0.5] | 0.709 |

\*median [inter-quartiles]; \*\*Low Dose Naltrexone; \*\*\*Post Exertional Malaise

**Table 2.** Symptoms and Functional Status Scale before and after initiation of Low Dose Naltrexone.

|  | **After LDN** | **Before LDN** | **P-value** |
| --- | --- | --- | --- |
| Number of symptoms | 13.5 [9, 17] * | 12 [6.75, 14.25] | 0.022 |
| Stage | 1.7% I<br>10% II<br>41.7% III<br>38.3% IV<br>8.3% V | 0% I<br>0% II<br>38.3% III<br>55% IV<br>6.7% V | 0.052 |
| Fatigue | 4 [3, 4] | 4 [3, 5] | 0.014 |
| Brain fog | 3 [2, 4] | 4 [2, 4] | 0.050 |
| Post exertional malaise | 4 [2, 5] | 4 [3.75, 5] | 0.033 |
| Unrefreshing sleep | 3 [2, 5] | 4 [3, 5] | 0.018 |
| Abnormal sleep pattern | 2.5 [0, 4] | 4[1.75, 4.25] | 0.010 |
| Lightheadedness | 2 [0, 3] | 2 [0, 3] | 0.455 |
| Headache | 3 [0, 4] | 3 [2, 4] | 0.036 |

\*Median [inter-quartiles]

When we looked at the impact of LDN on clinical symptoms in the aggregate, we found a significant benefit in reducing the total number of symptoms, functional status score, and selected symptoms (fatigue, brain fog, post-exertional malaise, unrefreshing sleep, abnormal sleep pattern, and headache). (Table 3, Figure 1-Box plot). However, there were no correlations between LDN dose and scored responses to the selected symptoms (correlation coefficient: headache 0.1809, fatigue -0.1013, brain fog -0.0717, unrefreshed sleep -0.1285, abnormal sleep pattern -0.1285, post-exertional malaise -0.1350 and lightheadedness -0.0045) (Supplemental material scatter plot graph).

**Figure 1.**
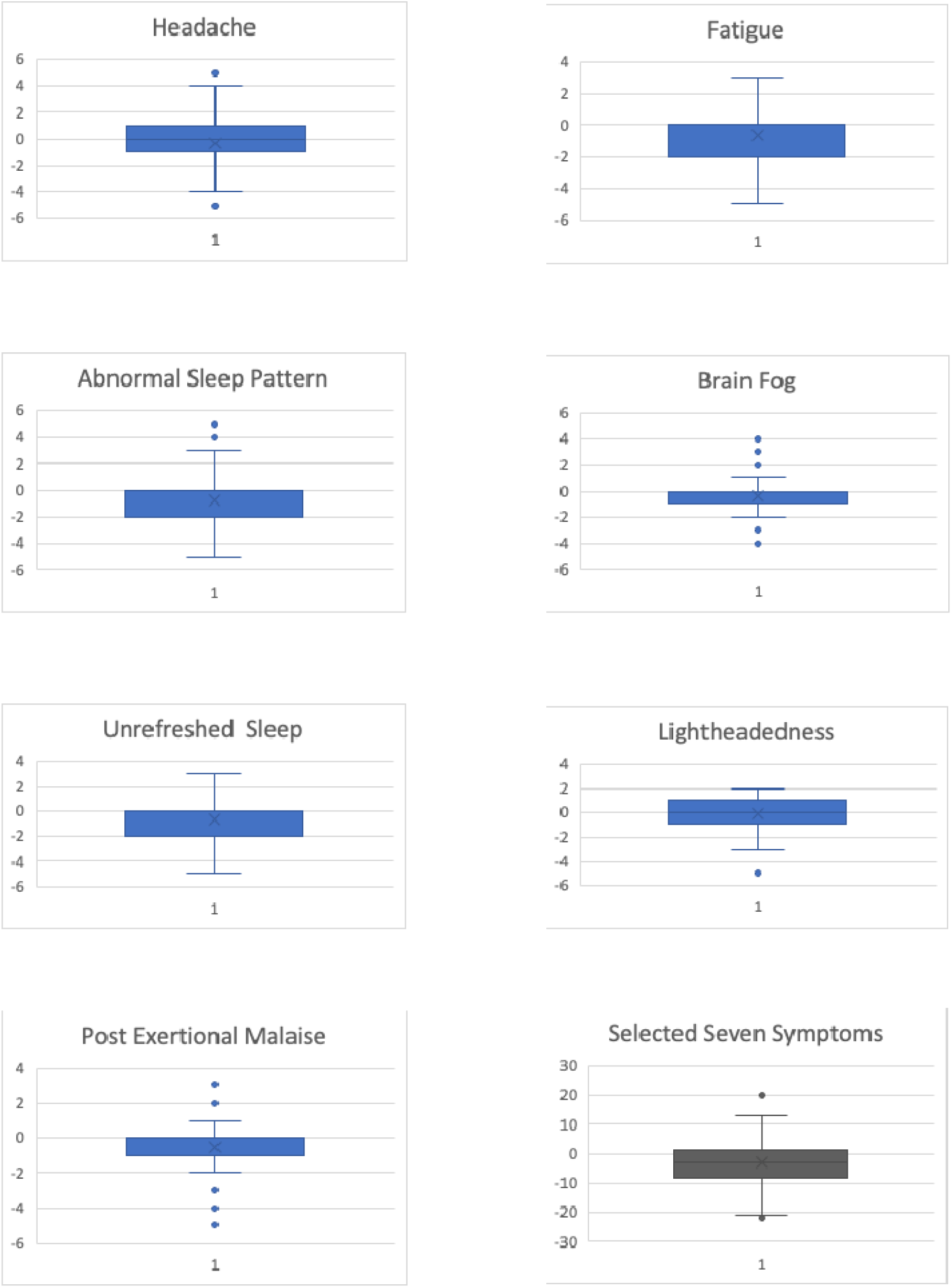
Plot box interquartile of the Likert score changes the severity of each seven (blue) and all selected symptoms (black) after a Low Dose of Naltrexone. Positive means worsening, and negative decrease severity.

## Discussion

The overall global prevalence of PASC is estimated at 43% of acute cases (54% among hospitalized 54% and 36% among non-hospitalized 36%)(9). The CDC estimates that 7.5% (over 24 million people) of adults have long COVID symptoms, and three times higher among 50–59-year-olds than individuals over 80 (12). The most prevalent symptoms include fatigue/weakness, myalgia/arthralgia, depression, anxiety, memory loss, concentration difficulties, dyspnea, and insomnia (9). Although the underlying biological mechanisms remain elusive, chronic inflammation driving end-organ disease is one of the main proposed hypotheses (3, 4, 13). There are currently no FDA-approved therapies for treating PASC symptoms, and there is an urgent need to find effective interventions. Naltrexone, an opioid receptor blocker drug with anti-inflammatory properties, is a promising candidate. The proposed anti-inflammatory and immunomodulator mechanisms of naltrexone are through attenuation of proinflammatory cytokines (IL-1, IL-6, TNF-α, and INF-β), nitric oxide, suppressive effects on microglia cells, Toll-like receptor-4 antagonist, and regulatory effects on NK cell function (14-17).

In our study cohort of 59 patients who received LDN as part of the off-label therapy for PASC, LDN use was associated with improvement in the primary symptoms explored in our study (fatigue, brain fog, PEM, unrefreshing sleep, abnormal sleep pattern, and headache). However, reported self-perception of improvement can introduce a bias when assessing the benefit of LDN. Although most patients with acute SARS-CoV-2 infection recover, in some patients, their symptoms can persist for months or even years. However, most of the recovery is a time-dependent variable. To mitigate the time factor on recovery in our study population, we compared the effect of LDN in individuals with ≥365 days of symptoms to those with <365 days of symptoms. In this PASC cohort, the duration of symptoms did not affect the clinical response to LDN to the selected symptoms. This retrospective EHR review of patients seen in our PASC clinic who received off-label use of LDN suggests a potential benefit of LDN in some individuals that warrants further testing in rigorous, randomized, placebo-controlled clinical trials. Given the heterogeneity of PASC, it will be important to select the suitable patient subpopulation that may best respond to LDN. In a cohort of patients with ME/CFS, the use of LDN showed clinical benefit in 73.9% (6). Most patients experienced improved vigilance/alertness and physical and cognitive performance. Some patients even reported less pain and fever. No severe adverse effects nor long-term adverse symptoms were reported (6). ME/CFS is characterized by debilitating fatigue and exertional intolerance, clinical features that overlap with PASC, and accumulating studies indicate about half of the patients with PASC have the ME/CFS phenotype (10).

A recently published open-label study evaluated the use of LDN in a cohort of 38 patients with long COVID without a control group. Six evaluated parameters improved over time in the 36 patients who completed two months of treatment, with the most significant reduction in pain (18). In contrast, in our study, the patients reported improvement in fatigue, brain fog, sleep, pain, and others (headache, anxiety, breathing, stuttering, and voice strength), and five patients had severe side effects such as: “nightmares,” “feeling poisoned,” and “worsening symptoms” that led to discontinuation of the drug. A larger study of LDN in 160 participants in a randomized parallel group double-blinded placebo-controlled trial is underway (18).

Finally, we did not find a correlation between LDN dose and changes in selected clinical symptom scores. This finding could be explained by multiple mechanistic pathways associated with long COVID (4). However, we cannot exclude the placebo effects that could go in either positive or negative response (19). Therefore, a randomized placebo-controlled trial will be necessary to mitigate this effect.

Our study is thus the most extensive observational study of LDN in PASC. However, our study has several limitations. First, this is a retrospective, observational, real-world setting without a placebo control or randomization. In addition, this study represents the experience of a single center in Northern California located in a generally affluent area with a bias toward specific populations. The selection of this cohort may skew our population in the following ways, 1) this is a referred population with multiple and more severe symptoms, and 2) our clinics have a lower proportion of underrepresented minority populations such as blacks and Hispanics, which lessen the generalizability of our findings. Finally, we did not assess the impact of other medications and supplements that patients were taking while on LND. Therefore, a multicenter study with a more diverse and larger population including several PASC phenotypes is necessary to corroborate our findings.

In summary, we describe a cohort of 59 patients with PASC who received LDN off-label as a potential therapeutic intervention for PASC. Overall, the use of LDN was associated with improved clinical symptoms of fatigue, brain fog, PEM, unrefreshing sleep, sleep pattern, and headache, and a reduced total number of symptoms with better functional status. Overall, these results suggest that LDN may improve symptoms of PASC and warrants further investigation in a randomized clinical trial. In addition, exploring the mechanism of action for naltrexone in neuroinflammatory conditions may also provide new insight into the pathogenesis of PASC.

## Supporting information

LDN Data

## Data Availability

It will be included as a supplementary material.

## Acknowledgements

We would like to thank the Stanford Health Care, and Stanford Department of Medicine for the clinic support, all patients, participants enrolled in the Stanford Long COVID clinic.

## Authors contributions

**Hector Bonilla**: (1) the conception and design of the study, acquisition of data, analysis and interpretation of data, (2) drafting the article, and revising it critically for important intellectual content, (3) final approval of the version to be submitted.

**Vincent Marconi**: (1) the conception and design of the study, analysis, and interpretation of data, (2) drafting the article, and revising it critically for important intellectual content, (3) final approval of the version to be submitted.

**Lu Tian**: (1) the conception and design of the study, acquisition of data, analysis and interpretation of data, (2) drafting the article, and revising it critically for important intellectual content, (3) final approval of the version to be submitted.

**Robert Shafer**: (1) the conception and design of the study, and analysis and interpretation of data, (2) drafting the article, and revising it critically for important intellectual content, (3) final approval of the version to be submitted.

**Grace A. McComsey**: (1) the conception and design of the study, analysis, and interpretation of data, (2) drafting the article, and revising it critically for important intellectual content, (3) final approval of the version to be submitted.

**Linda Geng**: (1) the conception and design of the study, analysis and interpretation of data, (2) drafting the article, and revising it critically for important intellectual content, (3) final approval of the version to be submitted.

